# The novel cardiokine GDF3 predicts adverse fibrotic remodeling post-myocardial infarction

**DOI:** 10.1101/2021.05.31.21257816

**Authors:** Nihar Masurkar, Marion Bouvet, Damien Logeart, Olivier Claude, Maguelonne Roux, Clément Delacroix, Damien Bergerot, Jean-Jacques Mercadier, Marc Sirol, Barnabas Gellen, Marine Livrozet, Antoine Fayol, Estelle Robidel, David-Alexandre Trégouët, Giovanna Marazzi, David Sassoon, Mariana Valente, Jean-Sébastien Hulot

## Abstract

**Background:** Myocardial infarction (MI) induces a repair response that ultimately generates a stable fibrotic scar. Although the scar prevents cardiac rupture, an excessive profibrotic response impairs optimal recovery.

**Objective:** To explore the regulation of fibroblasts proliferation through a paracrine action of cardiac stromal cells post-MI

**Methods:** We carried out a bioinformatic secretome analysis of cardiac stromal PW1^+^ cells isolated from normal and post-MI mouse hearts to identify novel secreted proteins. Functional assays were used to screen secreted proteins that promote fibroblast proliferation. The expressions of secreted proteins candidates were subsequently analyzed in mouse and human hearts and plasmas. The relation between levels of circulating protein candidates and adverse post-MI cardiac remodeling was examined in a cohort of 80 patients with a first ST-elevation MI and serial cardiac magnetic resonance imaging (MRI) evaluations.

**Results:** Cardiac stromal PW1^+^ cells undergo a change in paracrine behavior post-MI and secrete factors that promote fibroblast proliferation. Among these factors, growth differentiation factor 3 (GDF3), a member of the transforming growth factor-β family, was markedly upregulated in the ischemic hearts and induced fibroblast proliferation at high level. In humans, GDF3 was detected in the plasma at day 4 post-MI and GDF3 circulating levels were significantly associated with an increased risk of adverse remodeling 6-month post-MI (adjusted Odds Ratio (OR) = 1.76 [1.03 - 3.00], p = 0.037).

**Conclusions:** Our findings define a mechanism for the pro-fibrotic action of cardiac stromal cells through secreted cardiokines, such as GDF3, a candidate marker of adverse fibrotic remodeling following MI.

## Introduction

Acute myocardial infarction (MI) is characterized by the loss of cardiac muscle and its repair by a collagen-based scar (1). The development of an organized fibrotic scar after MI is a key process to replace the necrotic tissue and avoid cardiac rupture (2), however, a sustained profibrotic response leads to myocardial damage by excessively promoting the development of non-contractile fibrotic areas (2–4). The fibrotic process initiates immediately after the initial acute injury and involves proliferation and activation of fibroblasts to generate scar tissue (5). The level of cardiac fibrosis is an important predictor of clinical outcomes and a more precise assessment of the amplitude of the early profibrotic response might help in risk stratification post-MI (2,4,6,7). While serum biomarkers indicative of necrosis (cardiac troponin) and cardiac dysfunction (brain natriuretic peptide) have been identified, early markers of severe fibrosis development post-MI are less established and the identification of patients at high risk for developing large adverse fibrotic remodeling and heart failure remains challenging (2,8).

The cells that give rise to cardiac fibrosis are diverse and incompletely characterized (5,9). It is recognized that the post-MI scarring process involves fibroblasts but also other cell sources including cardiac stromal cells(9). Using PW1, a marker of multiple progenitor lineages (10,11), we identified a novel cardiac stromal cell sub-population which acquires a pro-fibrotic behavior following MI (12,13). PW1^+^ cells contribute directly to cardiac fibrosis to generate cells with a fibrogenic signature (12) and via interaction with the transforming growth factor-beta (TGF-β) pathways (13). Cardiac PW1^+^ cells are present in the close vicinity to resident fibroblasts following MI (12). In this study, we addressed whether PW1^+^ cells play a paracrine role that stimulates the proliferation of resident fibroblasts post-MI. We hypothesized that the paracrine behavior of cardiac PW1^+^ cells increases in response to MI and that the identification of these signaling molecules can serve as relevant biomarkers of the early profibrotic response as well as have predictive value in determining the degree of scar formation after MI.

## METHODS

Details on the procedures are provided in the online-only data supplement.

### Mouse MI model

All procedures and animal care protocols were approved by our institutional research committee (CEEA34 and French ministry of research, N° 2019050221153452) and conformed the animal care guideline in Directive 2010/63/EU European Parliament. All animals received humane care in compliance with the “Principles of Laboratory Animal Care” formulated by the National Society for Medical Research and the “Guide for the Care and Use of Laboratory Animals” prepared by the Institute of Laboratory Animal Resources and published by the National Institutes of Health (NIH Publication No. 86-23, revised 1996). Ischemic cardiac injury was performed in adult PW1^nLacZ^ reporter mice by permanent left anterior descending coronary artery (LAD) ligation, as previously described(12,13). Left anterior descending artery (LAD) permanent ligation was performed on male 8- or 13-week-old male C57BL/6J and PW1-reporter (PW1^nLacZ^) mice, which were anesthetized in an induction chamber with 2% isoflurane mixed with 1.0 L/min 100% O_2_ and placed on a supine position on a heating pad to maintain body temperature. Mice were intubated with endotracheal tube connected to a rodent ventilator (180 breaths/min and a tidal volume of 200 µL). During surgical procedure, anesthesia was maintained at 1.5-2% isoflurane with O_2._ The heart was accessed via an intercostal incision through the left side of the chest, and a pericardial incision. The LAD was exposed and encircled with an 8.0 prolene suture at the proximal position. The suture was briefly snared to confirm the ligation by blanching the arterial region. A similar procedure was followed for SHAM mice except that the thread around the LAD was not ligated. Mice were analyzed 7 days after LAD permanent ligation. Blood samples were collected in heparin-coated Eppendorf tubes and immediately centrifuged at 200 ×*g* for 15 min at 4°C to separate the plasma, which was stored at −80°C until analysis. Hearts were excised and immediately digested for FACS sorting or qPCR analysis.

### Cell isolation and fluorescence-activated cell sorting (FACS)

PW1^+^ cardiac cell sorting was performed as previously described (13). Small cell suspensions were prepared from total heart upon atria removal from 8-week-old PW1^nLacZ^ mice. The ventricles were enzymatically digested with collagenase II and dissociated. The following antibodies were used for cell sorting: BUV737-tagged anti-CD31 (1:100 dilution; BD Bioscience), BUV395-tagged anti-TER119 (1:50 dilution, BD Biosciences), phycoerythrin-cyanin7-tagged anti-CD45 (1:500 dilution; eBiosciences). To detect β-gal reporter activity, cells were incubated with the fluorescent substrate 5-dodecanoylaminofluorescein di-β-D-galactopyranoside (C_12_FDG) at 37°C for 1 h. The different populations were gated, analyzed, and sorted on a FACS Aria II cytometer (BD Biosciences).

### CyQUANT^™^ Cell Proliferation Assay

FACS-sorted PW1^+^ and PW1^−^ (FDG^−^) cells were seeded in 24-well plates at a density of 15,000 cells/well and cultured under normal conditions in Dulbecco’s modified Eagle’s medium (DMEM) supplemented with 10% fetal bovine serum (FBS, Sigma) and 1% penicillin and streptomycin (Sigma) for 5 days. The medium was collected and used to incubate serum-starved MEFs (cultured for 24 h under normal conditions and then serum starved for 24 h) for 24 h. The proliferation of MEFs of adult cardiac fibroblasts was evaluated using the CyQUANT^TM^ cell proliferation assay as per the manufacturer’s instructions. MEFs or adult cardiac fibroblasts incubated with complete medium served as the control. A similar protocol was used to tested the conditioned media collected 48 h after transfection of HEK293 cells.

### RNA-sequencing and bioinformatics analysis

We used RNA sequencing of freshly isolated PW1^+^ cells from normal and ischemic hearts as previously reported(13). In total, 300 ng of total RNA extracted from freshly isolated cells with SureSelect Strand-Specific RNA kit (Agilent) was used to prepare a library, according to the manufacturer’s instructions. The resulting library was quality checked and quantified by peak integration on Bioanalyzer High sensitivity DNA labchip (Agilent). A pool of equal quantity of 12 purified libraries was prepared, and each library was tagged with a different index. The mRNA pool libraries were finally sequenced on Illumina Hiseq 1500 instrument using a rapid flowcell. The pool was loaded on two lanes of the flowcell. A paired-end sequencing of 2× 100 bp was performed. After discarding reads that did not pass the Illumina filters and trimming low-quality sequenced bases (q < 28) using the Cutadapt program(14), we restricted our downstream analyses to reads with lengths greater than 90 bp. Selected reads were mapped to a murine reference transcriptome that was generated by the RSEM package(15) from the full mouse reference genome and the gtf transcript annotations file from ENSEMBL(16). Alignment and estimation of transcripts abundance in each of the 12 processed samples were performed using the RSEM program. Transcripts with abundance counts higher than 10 in more than two samples (N = 36,948) were considered as expressed and retained for further analysis. Abundances of transcripts assigned to the same gene were combined together, leading to the profiling of 16,403 genes. Analyses were conducted under the R environment (version 3.2.2). Galaxy 15.10 instance was locally installed on a server machine. WolfPsort, TMHMM, and SignalP were obtained from CBS prediction servers (https://services.healthtech.dtu.dk/, accessed April 26^th^, 2021). NLStradamus and PredictNLS were used in parallel to determine nuclear localization signals. Each dataset from RNA-seq, corresponding to a different population, was then processed through a pipeline designed to select sequences containing a signal peptide, no transmembrane segment, no nuclear localization signal, and structural features consistent with active secretion via classical or non-classical secretory pathways. This selection process allowed the identification of putatively secreted proteins as previously reported(17).

### HEK293 cell culture and transfection

HEK293 cells were cultured at 37°C in the presence of 5% CO_2_ in DMEM (Life Technologies) supplemented into 10% FBS and 1% penicillin and streptomycin. HEK293 cells were plated at 600,000 cells well in 6 well-plates in a medium without antibiotics. After 24 h, transfection of expression plasmids (Origene and Genscript) was performed with Lipofectamine® 2000 (Life Technologies) according to the manufacturer’s protocol using 2 µg of plasmids and 6 µL of Lipofectamine® 2000 diluted in Opti-MEM (Life Technologies). The cells were cultured for 2 days and then serum starved for 8 h prior to the collection of conditioned media and centrifugation at 200 ×*g* for 10 min. Supernatants were stored at −80°C.

### qPCR analysis

RNA was extracted from cardiac cells isolated from MI and SHAM C57BL/6J mice 7 days after surgery using the RNAqueous Micro Kit (AM1931, Invitrogen), as per the manufacturer’s instructions. Then, 500 ng of extracted RNA was subjected to reverse transcription using the SuperScript IV VILO kit (11756050, Invitrogen) as per the manufacturer’s instructions. The resulting cDNA was subjected to qPCR using SYBR Select Master Mix (4472908, Applied Biosystems) on Quant Studio 3 Real-Time PCR system (Thermo Fisher) as per the following condition: 95°C for 10 min, 40 cycles of 95°C for 15 s and 60°C for 1 min, followed by 95°C for 10 s and 60°C for 1 min. The expression of target gene was analyzed using the −2^ΔΔCT^ method following normalization to *RPL13* expression. The primer pair used are listed in Supplemental Table 1.

### Isolation of mouse embryonic fibroblasts

Primary MEFs were isolated from 13.5-days post-coitus C57bL/6J mouse embryos. The pregnant females were euthanized by cervical dislocation and embryos were surgically excised and separated from maternal tissues and the yolk sac in ice-cold phosphate-buffered saline (PBS). Embryos were then decapitated and eviscerated (removal of the heart, spleen, liver and intestine). The bodies were washed in ice-cold PBS to remove blood before being finely minced in a Petri dish without PBS. Samples were incubated for 15 min at 37°C in the digestion solution (0.05% trypsin-EDTA solution [Life Technologies], 0.1 mg/mL DNase 1 [Sigma]). The suspension was allowed to settle. The supernatant was drawn off, mixed with MEF culture medium (DMEM 4.5 g/L D-glucose [Life Technologies], 10% FBS, 1% penicillin-streptomycin, 1% non-essential amino acid [Life Technologies]) and centrifuged for 5 min at 200 ×*g*. After centrifugation, the pellet containing MEFs was resuspended in MEF culture medium. The pellet from the tissue digestion was resuspended in the digestion solution and incubated for 15 min at 37°C. The cells were allowed to settle, the supernatant was drawn off and processed as previously described. The cells from the first and second digestion steps were pooled and then plated in Petri dishes. Each Petri dish received a volume of cell suspension equivalent to 1.5 embryos.

After 12 h, the culture medium was changed to remove non-adherent cells and debris. The MEFs were passaged upon reaching 80% confluence. MEFs were harvested by trypsinization, centrifuged, and resuspended in a freezing medium (DMEM 4.5 g/L D-glucose, 1% penicillin-streptomycin, 10% dimethyl sulfoxide). Primary MEFs were cultivated between passage 0 and 4.

### Western blot analysis

Proteins were extracted from frozen mouse heart tissues using a Dounce-Potter homogenizer into ice-cold radioimmunoprecipitation assay (RIPA) buffer (50 mM Tris pH 7.4, 150 mM sodium chloride, 1% IGEPAL CA-630, 50 mM deoxycholate, and 0.1% sodium dodecyl sulfate [SDS]) supplemented with 1% anti-proteases (Sigma-Aldrich), 1% anti-phosphatase inhibitors (Phosphatase Inhibitor Cocktail 2 and 3, Sigma-Aldrich), and 1 mM sodium orthovanadate. After 1 h incubation at 4°C, the homogenate was centrifuged for 15 min at 15,300 ×*g* and 4°C and the supernatant containing proteins was collected. Cardiac PW1^+^ cells from 22 mice and PW1^−^ cells from 16 mice were pooled, centrifuged at 500 ×*g* for 15 min at 4°C, and lysed in urea-thiourea buffer (5 M urea, 2 M thiourea, 50 mM dithiothreitol [DTT], and 0.1% SDS in PBS, pH 7.4). Proteins were extracted as described above. Protein concentrations for all samples were determined using a Bradford-based protein assay (Bio-Rad).

After isolation of cardiomyocytes and non-cardiomyocytes from the adult mouse hearts, as previously described^7^, the proteins were denatured for 10 min at 70°C before loading on a NuPAGE™ Novex® 4-12% Bis-Tris gel (Life Technologies). After 3 h electrophoresis, proteins were transferred onto nitrocellulose membranes using the Trans-Blot Turbo Transfer System (Bio-Rad) and stained with 0.1% Ponceau S (w/v in 5% acetic acid) to assess transfer quality and homogeneous loading. Membranes were blocked for 1 h in Tris-buffered saline with 0.1% Tween-20 (TBS-Tween) containing 5% skim milk with constant shaking and then incubated for overnight at 4°C with primary antibodies specific for GDF3 (1:1000 for tissue and 1:500 for plasma, Abcam) and FLAG (1:1000, Sigma-Aldrich) diluted in 5% skim milk/TBS-Tween. After washing, the membranes were incubated for 1 h at room temperature (23°C) with horseradish peroxidase (HRP)-conjugated secondary antibodies diluted in 5% skim milk/TBS-Tween. Membranes were then washed and incubated for 5 min with SuperSignal™ West Pico PLUS Chemiluminescent Substrate (Life Technologies) before imaging with the Chemidoc® XRS+ camera (Bio-Rad) and analysis using the Image Lab™ software.

### ELISA for GDF3 quantification

Mice GDF3 levels were measured by GDF3 sandwich ELISA assay (GenWay, GWB-KBBHW6) and human GDF3 levels were measured by Human GDF3 ELISA kit (Fine Test, EH3126) following the manufacturer’s instructions. Briefly, standards and diluted samples (1:16 for mice and 1:2 for humans in standard diluent) were added to an anti-GDF3 microplate (pre-coated plate with an antibody specific for GDF3) and incubated for 60 minutes (mice) or 90 minutes (humans) at 37°C. After removing standards and samples, wells were washed and a biotinylated GDF3 detector antibody was applied. The plate was incubated for 1 h at 37°C. Wells were washed and incubated at 37°C with an avidin-HRP conjugate for 30 min for mice samples and with HRP-Streptavidin conjugate (SABC) for 30 min for human samples. Finally, after extensive washing, the wells were incubated with the 3,5,3′,5′-tetramethylbenzidine (TMB) substrate for 15 min in the dark at 37°C. The blue color product from the oxidation of TMB substrate changed into yellow after reaction termination with the addition of stop solution and incubation at 37°C for 15 min. Absorbance at 450 nm was quantitatively proportional to the amount of GDF3 captured in well and measured using microplate reader.

### PREGICA cardiac MRI sub-study

We used banked plasma from 80 patients with a first STEMI and who were enrolled in the prospective PREGICA cardiac MRI sub-study (Predisposition Genetical in Cardiac Insufficiency, clinicaltrials.gov identifier <u>NCT01113268</u>). Details of the study have been described previously(18). Briefly, the study involved 6 cardiology centers in France and enrolled patients between 18 and 80 years old referred for a first STEMI between 2010 and 2017 and seen within the first 24h after symptom onset. STEMI was defined by the presence of ST-segment elevation on the ECG, significant rise of troponin (≥3-fold higher than the upper limit reference) and the presence of at least 3 akinetic LV segments on the initial trans-thoracic echocardiography. Patients were not included if they had permanent atrial fibrillation, a diagnosis of previous MI or a history of cardiac disease. All patients had coronary angiography and primary PCI in the first 24h. Cardiac MRI was performed using a 1.5-T unit at day 4±2 after hospital admission and at 6-month follow-up in a subset of patients defining the CMR substudy of the PREGICA cohort. A standardized MRI protocol was followed in all centers and images were centrally analyzed. Cine images were acquired using a breath-hold steady-state free-precession sequence in long-axis and short-axis views. A stack of short-axis slices covering from the atrioventricular ring to the apex was used to derive left ventricular (LV) volumes, and ejection fraction (EF). Ten minutes after intravenous injection of gadolinium-based contrast agent, late gadolinium enhancement (LGE) images were acquired using a breath-hold segmented T1-weighted inversion-recovery gradient-echo sequence in the same long-axis and short-axis views of cine images. LGE images were assessed for infarct size. Blood samples were drawn at the same time than cardiac MRI. GDF3 was quantified on available plasma drawn at day 4 (n=80) using Elisa assays. The study was approved by the institutional review board, and all patients provided written informed consent.

### Statistical analysis

#### Mouse and in vitro studies

The number of samples (*n*) used in each experiment is recorded in the text and figure legends. All experiments were performed independently at least twice. The data are expressed as mean ± standard deviation (SD). Quantitative variables were analyzed with one-way analysis of variance (ANOVA, Kruskal-Wallis test) and pair-wise comparisons with Dunnett’s post-hoc test for multiple comparisons. The Mann-Whitney U test was used for comparing continuous variables between two groups.

#### Analyses of PREGICA cardiac MRI sub-study

P-values were obtained from Chi-Square test statistics for binary variables and by using the Mann-Whitney U test for comparing continuous variables between two groups. The association between GDF3 levels and the likelihood of presenting adverse cardiac remodeling was assessed by linear regression models, with additional adjustment for age and sex.

All statistical analyses were performed using R (version 3.6.2). A value of P < 0.05 indicated statistical significance.

## RESULTS

### PW1^+^ cells from ischemic hearts release factors that promote fibroblast proliferation

Adult PW1^nLacZ^ reporter mice were subjected to ischemic cardiac injury by left anterior descending coronary artery (LAD) ligation, as previously described(12,13). Hearts were harvested from mice with MI as well as SHAM-operated mice at day 7 post injury, and PW1^+^ cells were isolated by fluorescence-activated cell sorting (FACS) (**Figure 1a**). Following cultivation for 5 days, the conditioned media from these cells was collected and used to culture mouse embryonic fibroblasts (MEFs) for 24 h (**Figure 1a**). The effect of the conditioned media on the proliferation of MEFs was then evaluated. We found a significant increase in the proliferation of MEFs incubated with the conditioned media from PW1^+^ cells isolated from ischemic hearts as compared with those treated with the conditioned media from PW1^+^ cells from SHAM operated hearts (**Figure 1b**). There was no significant increase in response to conditioned media from PW1^-^ cells (**Figure 1b**). These observations suggest that activated PW1^+^ cells from the ischemic heart secrete pro-proliferative factors.

**Figure 1.**
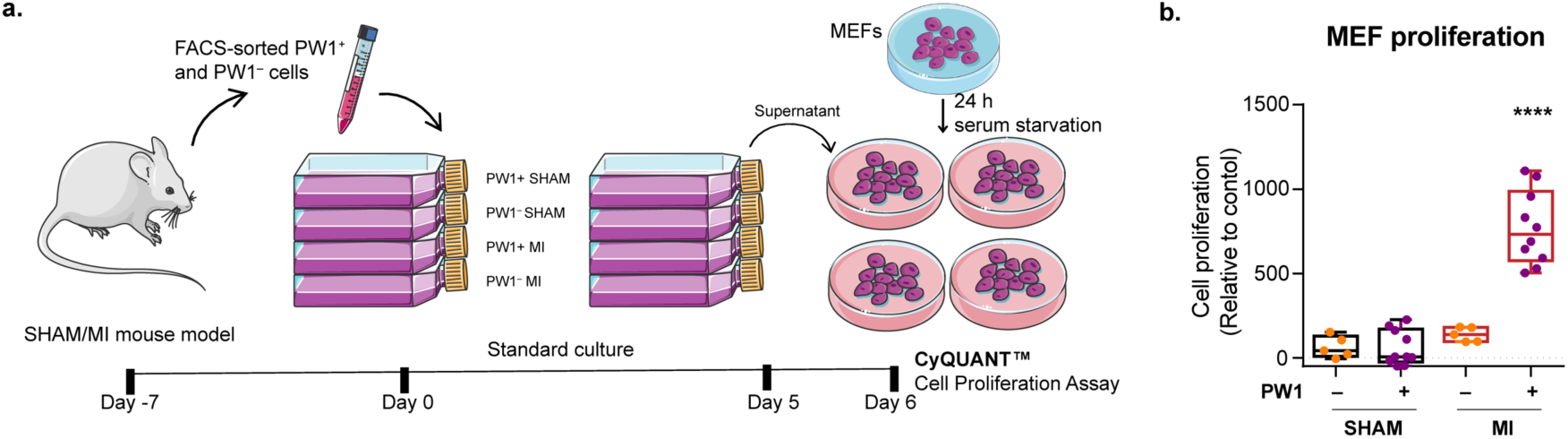
PW1^+^ cells from ischemic hearts release factors that promote fibroblast proliferation. **a.** Experimental workflow: FACS-sorted PW1^+^ and PW1^−^ (control) cells isolated from SHAM and MI mice after 7 days after surgery were cultured under standard conditions. On day 5, the culture supernatants were harvested and used to incubate serum-starved MEFs for 24 h. The proliferation of MEFs was evaluated using the CyQUANT™ cell proliferation assay. **b.** Comparison of proliferation of MEFs incubated with supernatants from different conditions. Statistical analysis was performed with one-way analysis of variance followed by post-hoc Dunnett’s multiple comparison test. ****P < 0.0001 versus PW1^+^ SHAM group. The experiment was performed three times in replicates of three.

### RNA-sequencing (RNA-seq) and bioinformatic analyses predict candidate biomarkers involved in the paracrine action

The transcriptome of FACS-isolated PW1^+^ cells from SHAM and MI mice was characterized by RNA-seq to investigate the influence of the ischemic heart environment on the paracrine potential of PW1^+^ cells. We filtered, aligned, and quality-controlled RNA-seq output files to obtain a list of transcripts showing the greatest signal intensities (**Figure 2a**). We first shortlisted candidates with more than two-fold higher expression in ischemic conditions than in normal condition (**Figure 2a**). We then examined the predicted amino acid sequences of the corresponding genes by a series of bioinformatic algorithms to identify secreted proteins. We considered proteins with a predicted N-terminal endoplasmic reticulum (ER)-targeting signal peptide but without predicted transmembrane domains and intracellular localization signals (i.e., no ER retention signal, mitochondrial targeting peptide, or nuclear export signal) (**Figure 2a**). We therefore defined a predicted secretome for each condition. Progressive filtering rendered a total of 24 candidates as secreted proteins overexpressed by cardiac PW1^+^ cells under ischemic conditions (**Figure 2a**). We next confirmed the significant increase in the expression of 12 of these 24 candidates in the ischemic heart (remote or infarct zones) as compared with normal heart by quantitative polymerase chain reaction (qPCR) (**Figure 2a-b and Online Figure 1**). In comparison with the secretome of control cells, the secretome of MI-activated PW1^+^ cells comprised several growth factors, cytokines, and enzymes as well as some poorly characterized factors (**Figure 2c**). The secretion of the growth differentiation factor-3 (GDF3), cytokines such as norrin cystine knot growth factor (NDP) and C-C Motif Chemokine Ligand 8 (CCL8), and enzymes such as chymotrypsin-like elastase family member 1 (CELA1) and proteinase 3 (PRTN3), were more than two-fold higher in MI hearts than in SHAM hearts (**Figure 2b,c**).

**Figure 2.**
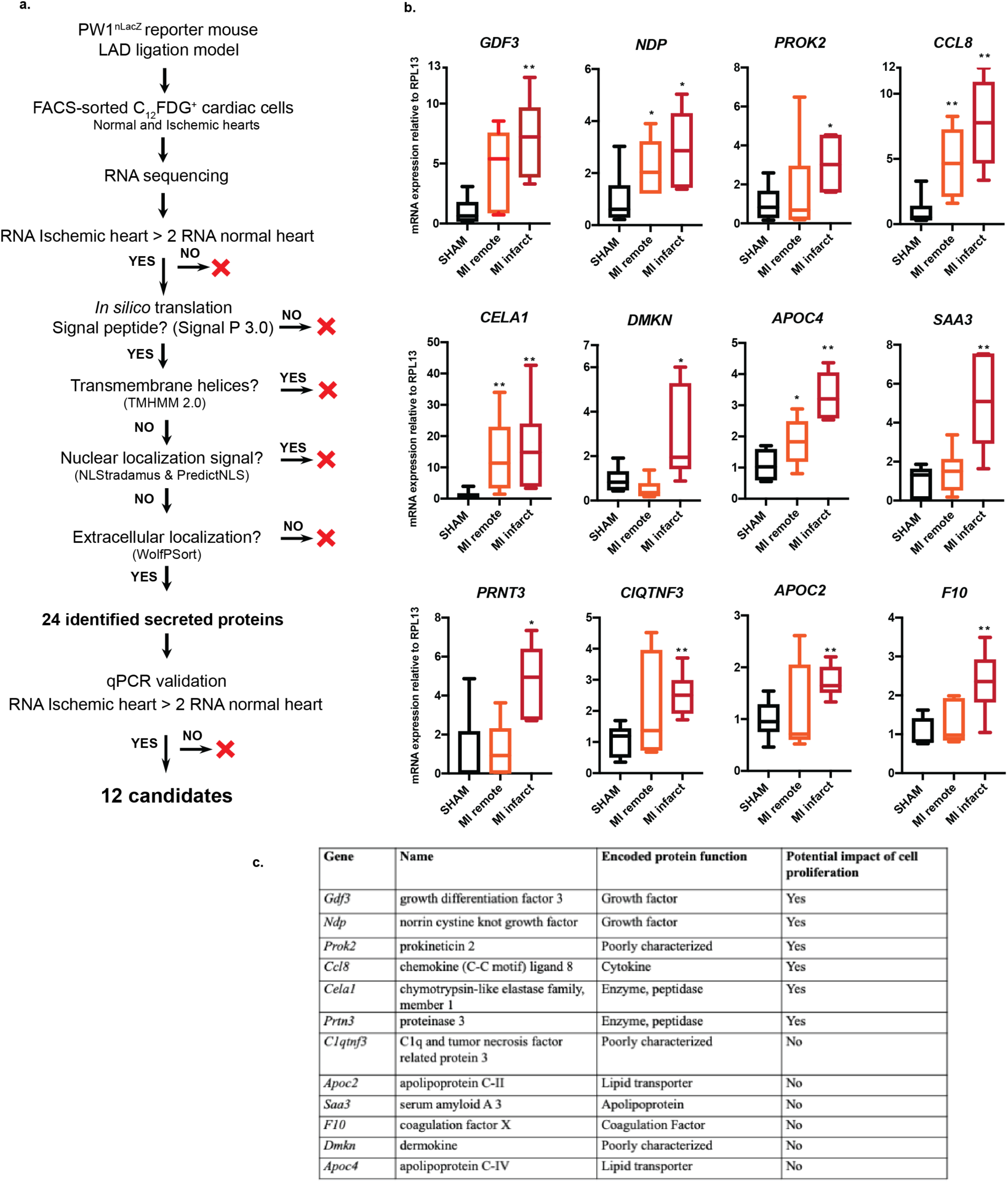
Prediction of candidate factors involved in the paracrine action of MI-activated PW1^+^ cells. **a.** Experimental strategy and bioinformatics algorithm to identify secreted proteins from FACS-purified PW1^+^ cardiac cells. **b.** The expression of 12 candidates was confirmed to be higher in ischemic than in normal hearts by RT-qPCR analysis. **P < 0.005 and *P < 0.05 for expression levels in infarcted or in remote areas versus levels in SHAM group, analyzed by Mann-Whitney tests; n = 6 each group. **c.** Names and known functional role of proteins encoded by each of 12 gene candidates.

### Identification of candidates with proliferative effects on fibroblasts

We next investigated the effects of candidates on the proliferation of cultured mouse embryonic fibroblasts and adult cardiac fibroblasts. We selected six candidates (CCL8, CELA1, GDF3, NDP, PRNT3, prokineticin 2 (PROK2)) that are associated with cell proliferation based upon assigned Gene Ontology biological functions. We excluded the lipoproteins APOC2, APOC4, and SAA3, the coagulation factor F10, and the poorly characterized C1QTNF3 and DMKN. We cloned cDNAs for these six candidate genes into mammalian expression plasmids, which were then used to transfect HEK-293 cells (**Figure 3a**). FLAG epitope-tagged fibroblast growth factor 23 cDNA served as positive control, while empty vector was used as negative control. Conditioned media were collected 48 h after transfection, tested to confirm the overexpression of the secreted proteins (**Figure 3b**), and then used to incubate serum-starved MEFs. Evaluation of cell proliferation rate after 24 h treatment revealed four factors, namely, GDF3, NDP, PROK2, and CELA1 that significantly induced the proliferation of MEFs as compared with control treatment (**Figure 3c**). The proliferative effects of GDF3, NDP and PROK2 but not CELA1 were further confirmed using freshly isolated adult cardiac fibroblasts (**Figure 3d**). Of these three remaining candidates, GDF3 appeared as the most promising candidate as combining both a high upregulation in the ischemic hearts (**Figure 2b**) and high induction of fibroblast proliferation (**Figure 3c-d**).

**Figure 3.**
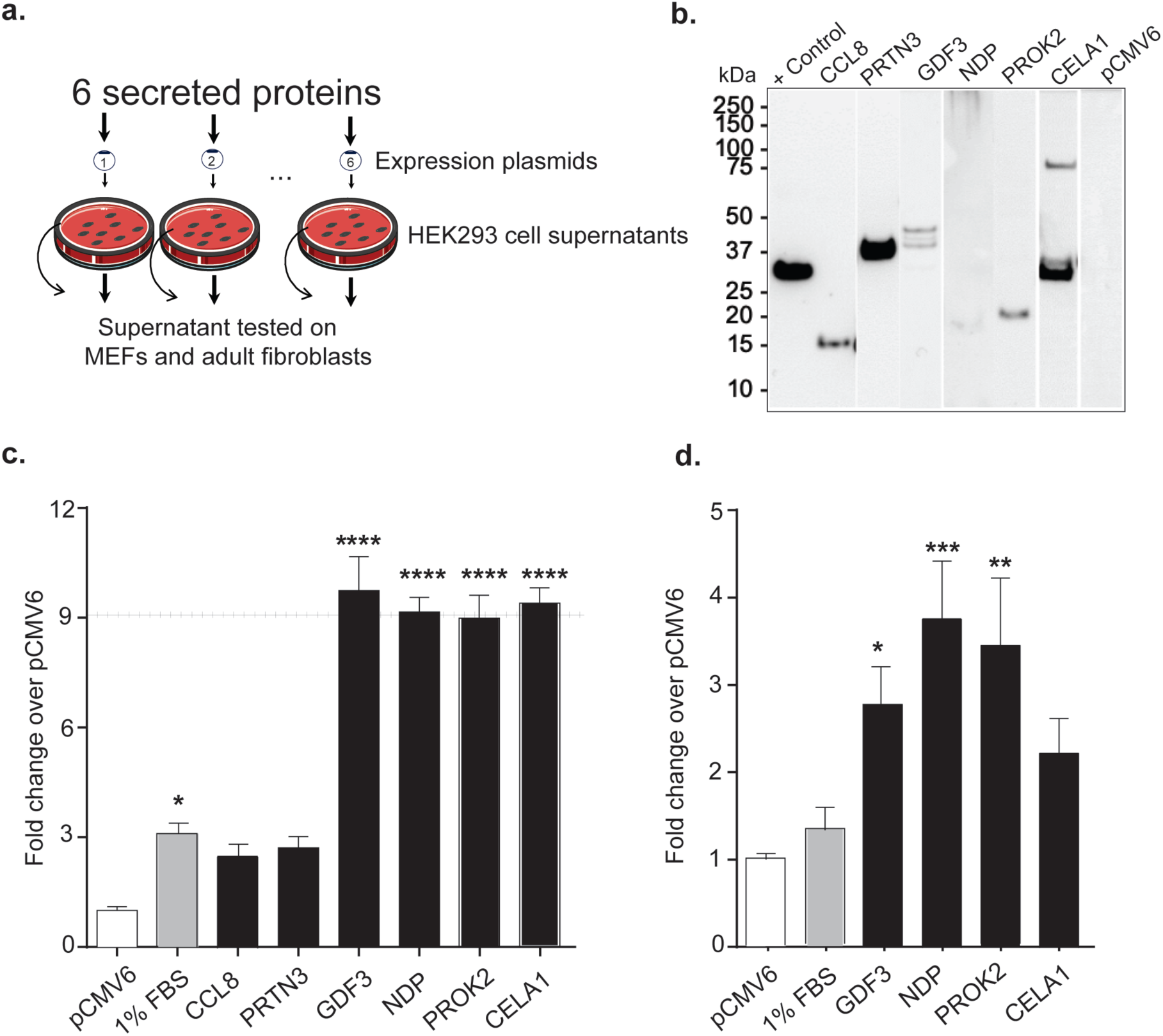
GDF3, NDP, and PROK2 induce MEF and adult cardiac fibroblast proliferation *in vitro*. **A.** Experimental strategy. Six candidate markers were cloned in an expression vector, which was used to transfect HEK293 cells cultured for 24 h. The cells were incubated for 48 h following transfection and then serum starved for 8 h. **b.** The supernatants collected were tested for the presence of candidate markers by Western blotting. **c. and d.** Plots of proliferation of MEFs (c) and adult cardiac fibroblasts (d) incubated with supernatants containing test candidates for 24 h. Statistical analysis was performed with one-way analysis of variance (ANOVA), followed post-hoc Dunnett’s multiple comparison tests. *P < 0.05, **P < 0.01, and ****P < 0.0001 versus the negative control pCMV6.

GDF3 (also known as Vg-related gene 2) is a member of the TGF-β superfamily that is composed of 366 amino acid residues. The predicted amino acid sequence comprises a signal sequence for secretion at the hydrophobic NH_2_-terminus, a prodomain that facilitates cysteine-mediated disulfide bond formation with another family member, and a putative proteolytic processing site at 114 amino acid residue (**Figure 4a**). The cleavage of GDF3 at this residue generates a mature GDF3 protein, which is 114 residue long (19) and human and mouse GDF3 show 76.6% nucleotide homology and 69.3% peptide identity. GDF3 plays an important role during early development in mice and humans (20), but its expression levels were predicted as typically low in the adult normal heart in gene expression atlases (21). While the function of GDF3 in the adult heart is unknown, GO biological functions suggest its involvement with the SMAD protein signal transduction pathway that is relevant to the process of cardiac fibrosis. These observations are consistent with the pronounced proliferative effect of GDF3 on MEFs (**Figure 3c**). Our transfection experiment (**Figure 3**) confirmed that GDF3 is a secreted protein, as indicated by the band of full-length protein on the Western blot of supernatants (**Figure 3b**). Taken together, these observations suggest a role for GDF3 in the regulation of fibroblast proliferation in the scarring tissue and prompted us to investigate the expression profile of GDF3 in MI hearts of mice and humans.

**Figure 4.**
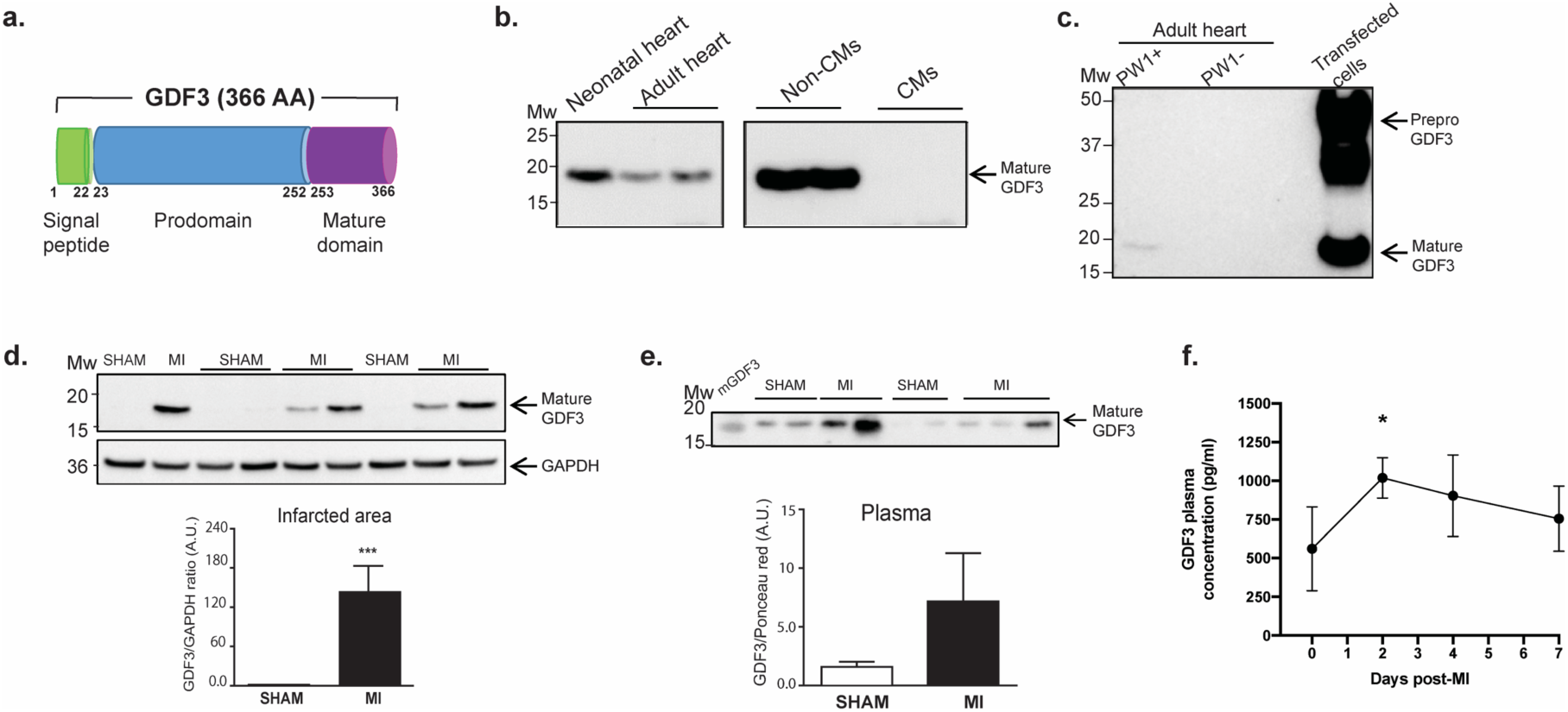
GDF3 expression is upregulated in the plasma and infarcted area of mouse heart following MI. **a.** GDF3 structure composed of a signal peptide, a prodomain, and a mature domain. **b.** Western blot analysis demonstrating the expression of mature GDF3 in the whole neonatal and adult mouse hearts (left panel) and in non-cardiomyocytes cell fraction (right panel). **c.** Within the adult normal heart, mature GDF3 is detected in FACS-sorted PW1^+^ cells but not PW1^−^ cells. **d.** Representative Western blots and quantification of mature GDF3 in the the infarcted area of MI mice (n = 9) compared to SHAM (n = 7). ***P < 0.001 by Mann-Whitney non-parametric t-test. **e.** Representative Western blots and quantification of secreted GDF3 in the plasma of SHAM (n = 7) and MI mice (n = 9) 7 days after MI. **f.** Kinetics of plasma GDF3 concentration in mice after MI (n ≥5 per time points). Quantification was performed by ELISA. * P < 0.05.

### GDF3 is upregulated in the infarcted area following MI

We evaluated the expression of GDF3 in normal and ischemic hearts by Western blotting. We detected the mature form of GDF3 in both neonatal and adult normal hearts (**Figure 4b, left panel**). In particular, GFD3 was expressed by the non-cardiomyocyte fraction and was undetectable in the cardiomyocytes of the adult heart (**Figure 4b, right panel**). Further analysis of the non-cardiomyocyte fraction confirmed the specific expression of GDF3 in PW1^+^ cells whereas it was undetectable in the PW1^−^ cell population of normal hearts (**Figure 4c**). Western blotting further confirmed that GDF3 was markedly up-regulated in the infarcted area of MI hearts 7 days after permanent LAD ligation as compared to the corresponding areas of SHAM hearts (**Figure 4d**). Consistent with our previous observations and with the fibrogenic fate of cardiac PW1^+^ cells in response to MI (12), this result indicates that GDF3 is produced at the site of infarction and suggests an involvement in the scarring process following MI.

### GDF3 is a circulating factor secreted post-MI

Considering the secretory nature of this protein, we investigated whether circulating GDF3 can be detected by analyzing plasma samples from MI and SHAM mice. Western blot analyses confirmed the presence of mature GDF3 in the plasma of MI mice 7 days after MI as compared to the plasma of SHAM mice (**Figure 4e**). We then performed an enzyme-linked immunosorbent assay (ELISA) specific for GDF3 to look for kinetic changes over time in the circulating levels of GDF3 in the mouse plasma after MI. The secreted protein level significantly increased from day 0 to day 2 and decreased thereafter until day 7 post-MI (**Figure 4f**). This result reveals the potential of circulating GDF3 as an early biomarker of post-MI remodeling in mouse.

### Circulating GDF3 level serves as a marker of post-MI adverse remodeling in humans

To investigate the clinical relevance of our findings in the murine MI model, we first assessed GDF3 expression on left ventricular cardiac tissue samples from failing ischemic hearts and non-failing hearts of patients. Western blot analyses revealed a marked expression of GDF3 in the failing ischemic hearts as compared to non-failing hearts (**Online Figure 2**), indicating the upregulation in the expression levels of GDF3 in the heart is a conserved response to MI. We then asked whether elevated circulating GDF3 levels can be linked to adverse cardiac remodeling post-MI. We analyzed circulating GDF3 levels in 80 patients with a first acute ST-elevation myocardial infarction (STEMI), seen < 24h after symptom onset and treated by primary percutaneous coronary intervention (PCI) (18). Patients had an initial clinical and biological evaluation at day 4 and serial cardiac magnetic resonance imaging (cMRI) at day 4±2 and at 6 months after angioplasty (**Figure 5a**). We defined adverse cardiac remodeling as a > 20% increase in left ventricular end-diastolic volume indexed for body surface area (LVEDVi, ml/m^2^) at 6 months as compared to the initial evaluation on cMRI. Patients were first classified as remodelers (n = 24) and non-remodelers (n = 56). The baseline characteristics of these patients are shown in Table 1.

**Figure 5.**
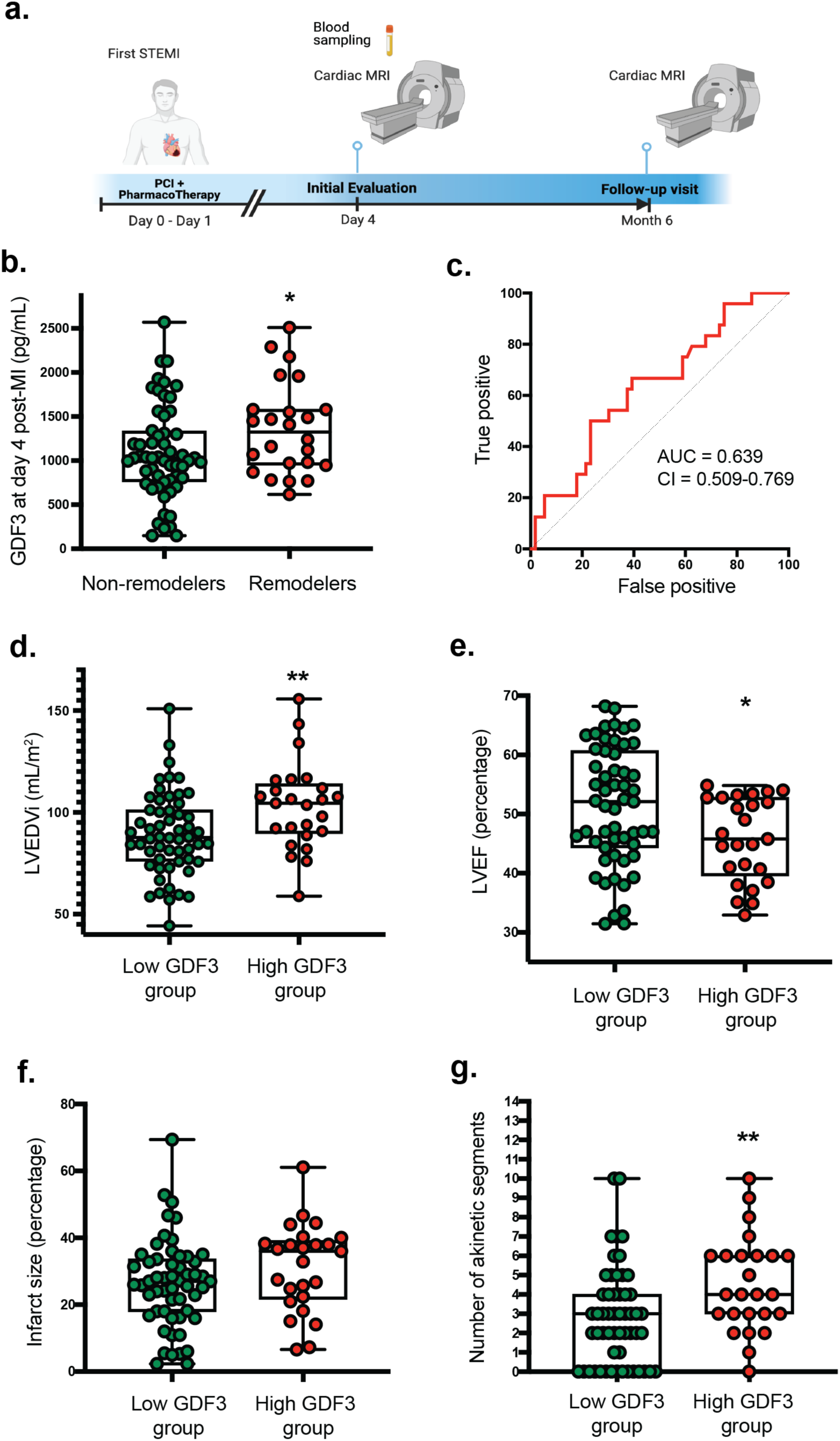
GDF3 is a circulating factor secreted post-MI and predicts adverse cardiac remodeling in humans. **a.** Experimental design of the PREGICA cardiac MRI sub-study **b.** GDF3 levels at day 4 post-MI in non-remodelers (n = 24) and remodelers (n = 56) as defined as a > 20% increase in LVEDVi at 6 months as compared to evaluation at day 4. *P = 0.05 as analyzed with Mann Whitney non-parametric t-test. **c.** ROC curve for the discrimination of GDF3 levels between remodelers and non-remodelers. **d-g.** LVEDVi, LVEF, infarct size, and number of akinetic segments at 6 months post-MI in patients from low GDF3 (<1375 pg/mL) and high (≥1375 pg/mL) GDF3 groups. *P < 0.05, **P < 0.01 by Mann Whitney non-parametric t-test.

**Table 1.** Characteristics of the overall population and according to cardiac remodeling at 6 months post-MI.

|  | **Overall**<br>**(N=80)** | **Non-**<br>**remodelers**<br>**(N = 56)** | **Remodelers**<br>**(N = 24)** | **Association**<br>**P-value** |
| --- | --- | --- | --- | --- |
| Age (years) | 58 ± 10 | 57 ± 10 | 60 ± 11 | 0.18 |
| Male, n (%) | 67 (83%) | 47 (84%) | 20 (83%) | 0.99 |
| BMI (kg/m<sup>2</sup>) | 25.9±4.1 | 25.9±3.8 | 26.1±4.8 | 0.83 |
| Pre-existing<br>hypertension, n (%) | 27 (33%) | 20 (36%) | 7 (29%) | 0.72 |
| Dyslipidemia, n<br>(%) | 47 (59%) | 37 (66%) | 10 (42%) | 0.07 |
| Diabetes, n (%) | 15 (19%) | 11 (20%) | 4 (17%) | 0.99 |
| Time between<br>symptoms onset<br>and PCI (hours) | 5.11 ± 4.30 | 5.08 ± 4.52 | 5.19 ± 3.80 | 0.91 |
| LAD as a culprit<br>artery, n (%) | 52 (65%) | 35 (62.5%) | 17 (71%) | 0.47 |
| T Troponin peak<br>(µg/L) | 99 ± 112 | 99 ± 122 | 101 ± 82 | 0.91 |
| CRP (mg/L) | 29.5 ± 48.6 | 22.7 ± 34.4 | 45.5 ± 70.3 | 0.37 |
| Hb1AC (%) | 6.13 ± 1.10 | 6.11 ± 1.12 | 6.18 ± 1.09 | 0.68 |
| GDF3 (pg/mL) | 1172 ± 540 | 1090 ± 532 | 1364.0 ± 521 | **0.03** |

Mean ± SD values are shown for continuous variables while count (%) are given for binary variables. LAD, left anterior descending artery; PCI, percutaneous coronary intervention; BMI, body mass index;

GDF3 measured at day 4 post-PCI was detectable in the plasma of these patients (**Online Figure 3**). Plasma GDF3 levels were significantly higher in remodelers than in non-remodelers (1350±521 vs 1090±532 pg/ml, p = 0.033) (**Figure 5b**). After adjusting for age and sex, one standard deviation (SD) increase of GDF3 levels was associated with an increased risk of adverse remodeling (Odds Ratio (OR) = 1.76 [1.03 - 3.00], p = 0.037). Plasma GDF3 levels did not demonstrate any statistical difference (p>0.10) according to sex, smoking, personal history of hypertension nor diabetes. Of note, GDF3 levels were moderately correlated to CRP (ρ = 0.13) and Hb1AC (ρ = 0.18), even if these correlations did not reach statistical significance (p = 0.28 and p = 0.12, respectively).

To better assess the relation between cardiac remodeling and plasma GDF3 levels, we first divided patients into 4 quartiles of GDF3 levels (measured at day 4 post-MI) and compared LVEDVi and LVEF measured on cardiac MRI 6 months after MI between the quartiles. We found that patients with the highest GDF3 levels (i.e., quartile 4) had significantly higher LVEDVi and lower LVEF as compared to patients with lower GDF3 levels (**Online Figures 4 and 5**). We next performed a receiver operating characteristic (ROC) curve analysis to determine if GDF3 level could help distinguishing the two groups. The area under the receiver operating characteristic curve of the age and sex adjusted model was 0.69 [0.56-0.82] (p = 0.05) and yielded a cut-off value of 1375 pg/mL at a likelihood ratio of 2.154 and sensitivity and specificity of 50% and 77%, respectively (**Figure 5c**). This cut-off value was close to the one defining quartile 4 (**Online Figures 4 and 5**) and was then used to classify patients into two groups (≥ 1375 pg/mL called high GDF3 -red-, n = 25 and < 1375 pg/mL called low GDF3 - green-, n = 55). Table 2 reports the main characteristics and cardiac MRI findings at baseline and 6 months after MI in both groups. There was no significant imbalance in the main cardiovascular risk factors between the two groups. The peak of troponin, a surrogate marker of myocardial necrosis, was significantly lower in the high GDF3 group. In terms of cardiac remodeling, patients with high GDF3 showed a non-significant trend of higher cardiac dilation at day 4 post-MI than those with low GDF3. The values of LVEDVi were however significantly higher (P < 0.005) and pathological (normal value < 82 mL/m^2^) at 6 months post-MI in patients with high GDF3 (**Figure 5d**), indicative of adverse cardiac remodeling along with the development of cardiac dilation. These patients also showed a significant decrease (p < 0.05) in LVEF at day 4 and 6 months (**Figure 5e**), suggesting the reduced recovery of contractile function post-MI in these patients. While total infarct size (percentage total cardiac mass) estimated on cardiac MRI was not significantly different between the two groups (**Figure 5f**), patients with high GDF3 levels had a higher proportion of akinetic segments on cardiac MRI at 6 months than those with low GDF3 levels (**Figure 5g**). This result indicates the more important pathological transformation of infarcted areas into non-contractile scars in patients with high GDF3 levels, emphasizing its diagnostic importance as a marker of adverse cardiac remodeling after MI.

**Table 2.** Main characteristics and cardiac MRI findings at baseline and 6 months after MI in both groups according to GDF3 cut-off.

| Characteristics | Patients with GDF3<br>< 1375 pg/mL<br>(N = 55) | Patients with<br>GDF3 ≥ 1375<br>pg/mL<br>(N = 25) | P value |
| --- | --- | --- | --- |
| Age (years) | 59 ± 9 | 56 ± 13 | 0.17 |
| Males, n (%) | 44 (80%) | 23 (92%) | 0.31 |
| BMI (kg/m<sup>2</sup>) | 25.7 ± 4.2 | 26.4 ± 3.8 | 0.21 |
| Pre-existing hypertension, n (%) | 21 (38.9%) | 6 (24%) | 0.06 |
| Dyslipidemia, n (%) | 36 (65%) | 11 (44%) | 0.07 |
| Diabetes, n (%) | 8 (14.5%) | 7 (28%) | 0.15 |
| Time between symptoms and PCI (hours) | 4.90 ± 4.48 | 5.47 ± 3.86 | 0.56 |
| LAD as a culprit artery, n (%) | 35 (63.6%) | 17 (68.0%) | 0.13 |
| ***Biology at day 4*** |  |  |  |
| T Troponin peak (μg/L) | 115 ± 117 | 66 ± 90 | **0.0052** |
| C-reactive protein (CRP) | 26.8 ± 40.2 | 35.9 ± 64.9 | 0.55 |
| GDF3 (pg/mL) | 875 ± 301 | 1826 ± 330 | **<0.0001** |
| ***Cardiac parameters (MRI) at day 4*** |  |  |  |
| LVEDVi (mL/m<sup>2</sup>) | 82.4 ± 16.6 | 91.8 ± 16.1 | 0.052 |
| LVESVi (mL/m<sup>2</sup>) | 43.9 ± 13.2 | 52.4 ± 12.9 | **0.0097** |
| LVEF (%) | 47.3 ± 8.4 | 43.3 ± 7.4 | **0.0296** |
| Number of akinetic segments | 4.66 ± 2.27 | 5.64 ± 2.15 | **0.035** |
| ***Cardiac parameters (MRI) at Month 6*** |  |  |  |
| LVEDVi (mL/m<sup>2</sup>) | 89.3 ± 20.2 | 103.0 ± 21.3 | **0.0065** |
| LVESVi (mL/m<sup>2</sup>) | 44.3 ± 17.1 | 56.5 ± 17.1 | **0.0032** |
| LVEF (%) | 51.3 ± 10.0 | 45.8 ± 7.1 | **0.0129** |
| Infarct size (%) | 26.3 ± 13.2 | 31.2 ± 13.0 | 0.08 |
| Number of akinetic segments | 2.80 ± 2.51 | 4.52 ± 2.47 | **0.0037** |

Mean±SD values are shown for continuous variables while count (%) are given for binary variables. LAD, left anterior descending artery; PCI, percutaneous coronary intervention; BMI, body mass index; LVEDV, left ventricular end-diastolic volume; LVESV, left ventricular end-systolic volume; LVEF, left ventricular ejection fraction.

## DISCUSSION

We showed previously that cardiac stromal PW1^+^ cells have a pro-fibrotic behavior in the context of MI via direct differentiation into fibroblasts-like cells (12) and interaction with the transforming growth factor-beta (TGF-β) pathways (13). We show here that cardiac PW1^+^ cells exert a pro-fibrotic paracrine effect on fibroblasts after MI. The paracrine activity of cardiac PW1^+^ cells is modulated in response to MI and includes an increased expression of cytokines and growth factors with pro-proliferative effects on fibroblasts. We identified candidate molecules underlying this paracrine effect by evaluating changes in the transcriptional profiles of cardiac PW1^+^ cells in response to MI and by using a bioinformatic suite to predict secreted proteins as previously described (17). Among these factors, we identified growth differentiation factor 3 (GDF3), which is a member of the TGF-β superfamily (19,22) and plays key role during embryonic development (20,23). GDF3 was shown to participate in muscular development (24), adipose tissue homeostasis(25), and energy balance through its interaction with the activin receptor-like kinase type I receptor B (ACVR1B, ALK4) and ACVR1C (ALK7) receptors(22,26). GDF3 is typically expressed at low levels in the normal adult heart (19,21). There is a limited knowledge on the biological function of GDF3 in adult hearts as well as on cardiac fibroblasts. Even if TGF-β pathways have been consistently highlighted as the key molecular mediators of cardiac fibrosis (13,27), this is the first report associating GDF3 with cardiac fibrosis. GDF3 belongs to the same protein subfamily than GDF15, another cardiokine that regulates leukocytes activation after MI (28) and has been proposed as a prognostic biomarker in a variety of cardiovascular diseases (29). However, the protein sequence of GDF3 has a limited homology with GDF15 (26% in humans, 33% in mice) and GDF3 cannot be disulfide-linked in contrast to other members of this subfamily suggesting a distinct structure and function.

We show here that the upregulation of GDF3, a secreted protein, is detected in the plasma of mice and humans following MI. We interpret that the levels of circulating GDF3 correlates with the local cardiac production in response to MI and that higher GDF3 circulating levels would indicate higher proliferation of fibroblasts and higher fibrogenesis. Concordantly, we show that high levels of plasma GDF3 levels in a cohort of post-MI patients 4 days following MI(18) corresponded with adverse outcomes measured 6 months later including cardiac dilation, limited recovery of contractile function, and higher number of akinetic segments. These data suggest that higher circulating GDF3 levels can be used to identify patients who will develop adverse cardiac remodeling. It is well known that acute myocardial infarction is characterized by left ventricular remodeling that can progress toward heart failure in some patients (30). Markers reflective of myocardial damage, such as troponin and creatine kinase, may not predict long-term left ventricular remodeling (2,31). Other circulating markers of myocardial fibrosis, including molecules related to collagen metabolism or to inflammation, have been reported but their associations with the extent of myocardial fibrosis is either inconclusive (31) or not specifically associated with the development of fibrotic scar post-MI. Risk stratification at an early stage after MI is challenging yet useful to tailor personalized treatment regimen in the future. There is currently a lack of early markers that could indicate patients with an increased risk of adverse cardiac remodeling (31). Future studies are now needed to evaluate the predictive value of GDF3 to predict patients who will develop ischemic heart failure. Last, our study paves a strong foundation for future studies directed to target GDF3 in the treatment of MI.

Cardiac extracellular matrix (ECM) undergoes constant remodeling upon injury(2,5,32). Interestingly, the concept of ECM regulation through key molecules involved in intercellular communications has only recently emerged. Here, we focused on cardiac stromal PW1^+^ cells, a cellular subpopulation that is suspected to orchestrate the reparative process in tissues, including the heart. Of note, the pro-proliferative effect observed with conditioned media from cardiac PW1^+^cells isolated from ischemic mouse hearts was not observed with cardiac PW1^-^ cells nor with cardiac PW1^+^cells isolated from normal mouse hearts. We confirmed the upregulation in the expression of 12 secreted factors by MI-activated cardiac PW1^+^ cells. These factors were mostly enriched in GO biological processes such as angiogenesis, inflammation, chemotaxis, and proliferation. This result indicates an unprecedented role for cardiac PW1^+^ cells as key paracrine signaling cells that drive the repair response to injury.

Our clinical study is limited by the small sample size, as we specifically focused on post-MI patients with cardiac MRI evaluation for cardiac remodeling, an investigation that is not routinely performed in these patients. Our results indicate that patients with high GDF3 levels develop adverse cardiac remodeling based on validated imaging surrogates (4), but further studies are required to validate its prediction on heart failure and cardiovascular outcomes. However, a large proportion of patients with high GDF3 levels had a significantly decreased LVEF (< 50%) 6 months after MI. Further, while we only focused on the potential implications of GDF3, the roles of other upregulated markers, particularly PROK2 and NDP, was not investigated and warrant future studies. Notably, we cannot exclude a potential synergy between these other identified secreted factors. Lastly, this study investigated the role of secreted factors on fibroblast proliferation as a primary mechanism underlying cardiac fibrosis. However, other mechanisms support the fibrotic transformation of the ischemic heart such as myofibroblast transformation(27) and immune-inflammatory response(32,33). A role for GDF3 in macrophage polarization and endotoxin/sepsis-induced cardiac injury was recently reported(34). MI is characterized by an acute inflammatory response involved in myocardial repair(32) and uncontrolled chronic inflammation causes excessive damage and fibrosis, eventually leading to the loss of cardiac function. The impact of GDF3 on these mechanisms merits further investigations.

## CONCLUSION

In conclusion, PW1^+^ cells from the ischemic heart release pro-proliferative factors following MI, which promote proliferation of resident fibroblasts. One such factor, GDF3 may serve as a novel marker of adverse fibrotic remodeling in the heart tissue following MI. Its applicability in the clinical setting may allow for the identification of patients that have an increased risk of severe myocardial fibrosis and HF as well as better and more specific disease management.

## Data Availability

on request

## Abbreviations

CELA1: chymotrypsin-like elastase family member 1
FACS: fluorescence-activated cell sorting
FDG: fluorescent substrate 5-dodecanoylaminofluorescein di-β-D-galactopyranoside
GDF3: growth differentiation factor-3
LAD: left anterior descending coronary artery LAD
LVEDVi: left ventricular end-diastolic volume indexed for body surface area
LVEF: left ventricular ejection fraction
NDP: norrin cystine knot growth factor
MEFs: mouse embryonic fibroblasts
MI: Myocardial Infarction
MRI: Magnetic resonance imaging
PROK2: prokineticin 2
STEMI: ST-elevation myocardial infarction
TGF-β: transforming growth factor-beta

## Acknowledgments

RNA sequencing was performed with the help of Plateforme P3S and cell sorting with the help of Plateforme CyPS at Sorbonne Universités, Paris, France or Flow cytometry platform at the Paris Cardiovascular Research Center (PARCC).

## Perspectives

### Competencies in medical knowledge

Factors are secreted by the heart after MI, and these cardiokines can exert pro-proliferative actions on local cardiac fibroblasts thereby promoting a sustained fibrotic response.

### Translational outlook

The levels of circulating GDF3, a novel identified cardiokine, measured in the days after MI can help predicting patient subgroup at a higher risk of further developing adverse fibrotic remodeling and subsequent heart failure.

---

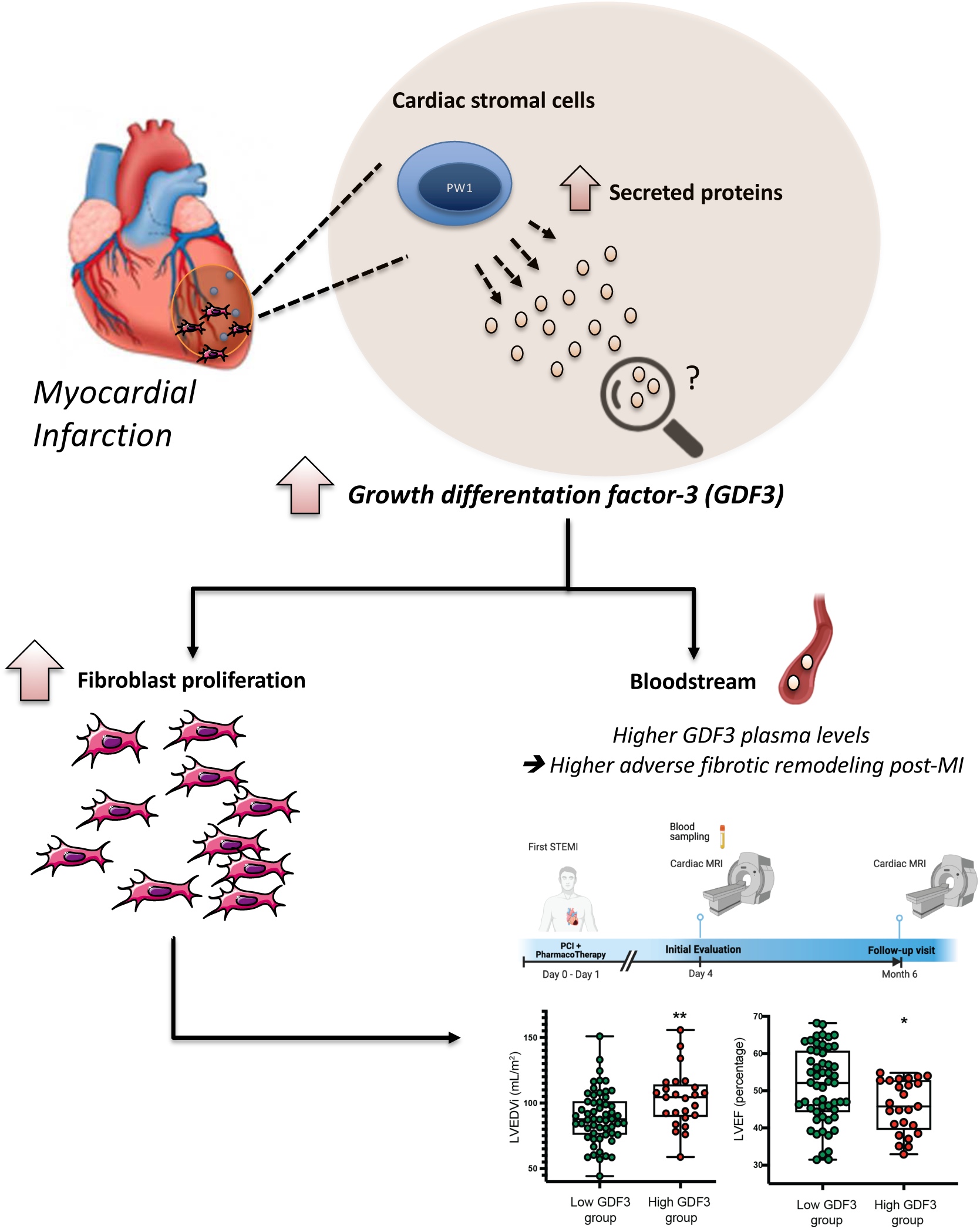
Central Illustration. Cardiac stromal PW1^+^ cells undergo a change in paracrine behavior post-MI and secrete factors that promote fibroblast proliferation. Among these pro-proliferative cardiokines, growth differentiation factor 3 (GDF3) is upregulated in the infarcted cardiac tissue and in plasma. In humans, the higher plasma GDF3 levels predict higher fibrotic cardiac remodeling and cardiac dilation 6 months post-MI.

