## supplemental tables and figures for "The novel cardiokine GDF3 predicts adverse fibrotic remodeling post-myocardial infarction"

### SUPPLEMENTAL MATERIAL

**Supplemental Tables : 1**

**Supplemental Figures : 5**

**Supplemental Table 1. PCR primers**

| GENE | FORWARD PRIMER (5' TO 3') | REVERSE PRIMER (5' TO 3') |
|---|---|---|
| ***CCL8*** | TCTACGCAGTGCTTCTTTGCC | AAGGGGGATCTTCAGCTTTAGTA |
| ***PRTN3*** | ATGGCTGGAAGCTACCCATC | TGCCCACCTACAATCTTGGAG |
| **F10** | AGGACTCGGAGGGCAAACT | TCACGGACCTCTTCATAAGAACA |
| **SAA3** | TGCCATCATTCTTTGCATCTTGA | CCGTGAACTTCTGAACAGCCT |
| **APOC4** | TGTTCTTGGTCAGCTTTGTAGC | AGGCTGTGGGTCTTGTTTAGG |
| **APOC2** | ATGGGGTCTCGGTTCTTCCT | GTCTTCTGGTACAGGTCTTTGG |
| **PROK2** | TTGCGACAAGGACTCTCAGT | CCCATAGGTCAGATCCT |
| **CIQTNF3** | GCTGGTAAACATAGGTGGCG | GCGGTTCTTCATCAGCTTCA |
| **CELA1** | GGGGCTCCTCTGTGAAGAAT | AGCATACTGACCGTTCACCA |
| **GDF3** | TTCAGCTTCTCCCAGACCAG | CCTTTTCTTTGATGGCAGACAAG |
| **DMKN** | CGGGAGTCACACCTTCATCT | AACTTCAGCCACTTCAGCAG |
| **NDP** | AGTCTGAGAAGAAGGAGCCC | GCTTCTTTCACTTGCAACCG |
| **RPL13** | GGGTGGCCAGCTTAAGTTCT | GAGGAGGCGAAACAAGTCCA |

**Online Figure 1. Expression of candidates that were not increased in the ischemic heart.** qPCR evaluation of *REG3G*, *F9*, *CFD*, *17007RIK*, *TRH*, *GM26947*, *ADIPOQ*, *PNOC*, *LIPC*, *MUC6*, *MZB1*, and *AMBP* expression in the infarcted area of MI hearts and the corresponding area of sham hearts (N = 3). Statistical significance was determined by Mann-Whitney test, *P < 0.05, **P < 0.01 and ***P < 0.001.

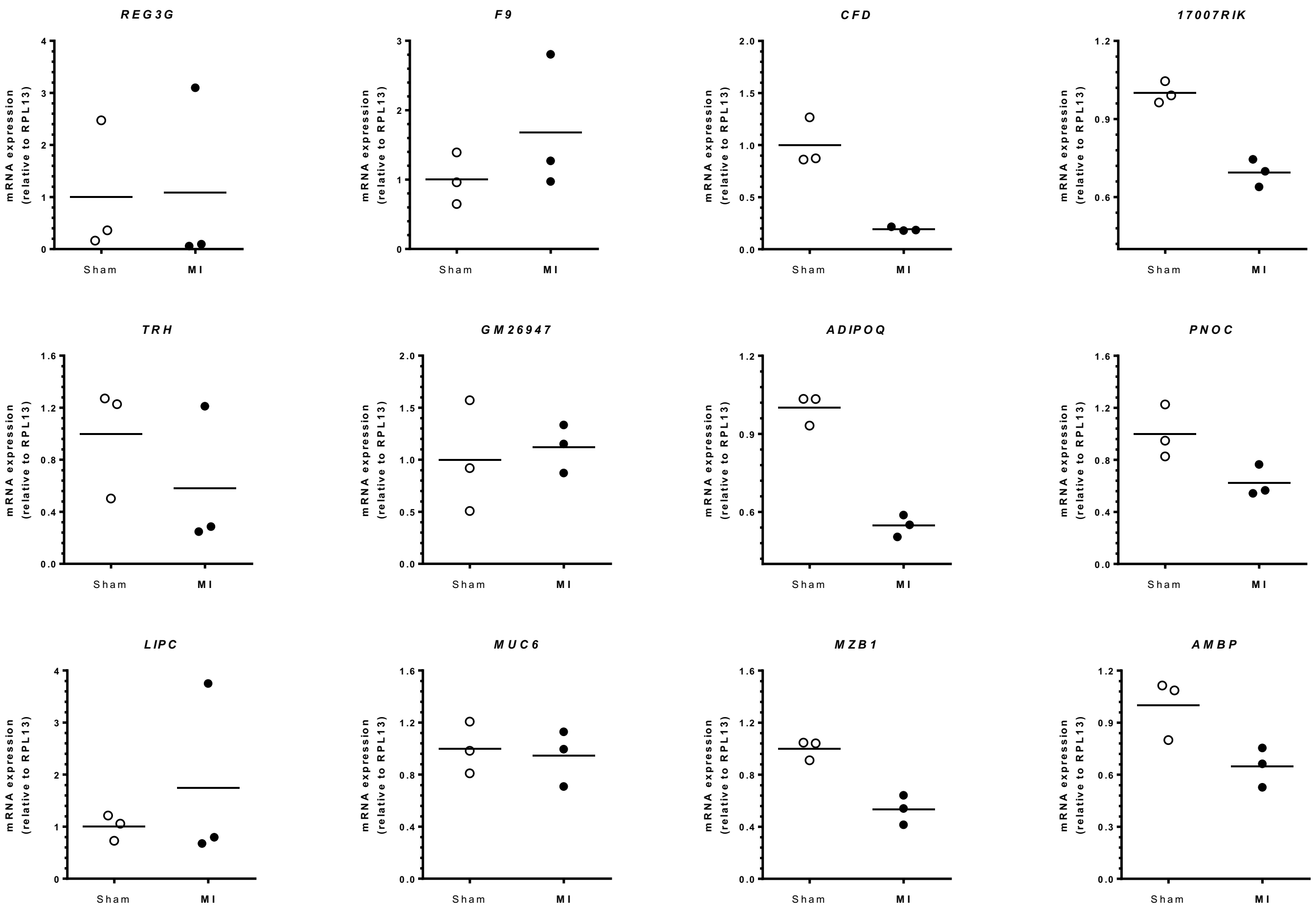

**Online Figure 2. Expression of GDF3 in human hearts.** Representative Western blots and quantification of mature GDF3 in non-failing (NF) (n = 6) and failing hearts (HF) (n = 9) of patients. *P < 0.05 as determined by Mann Whitney test

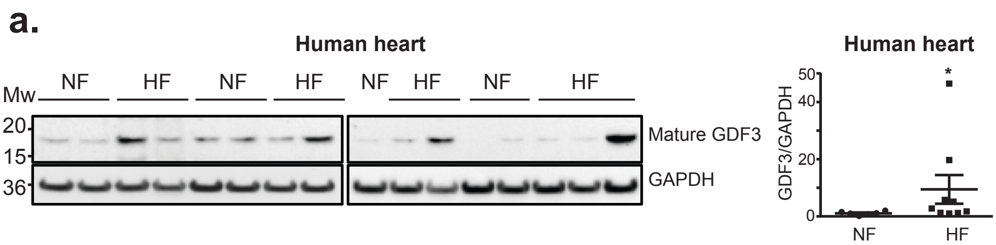

**Online Figure 3. Distribution of plasma GDF3 levels**. GDF3 was measured by ELISA in human plasmas collected at day 4±2 post-PCI.

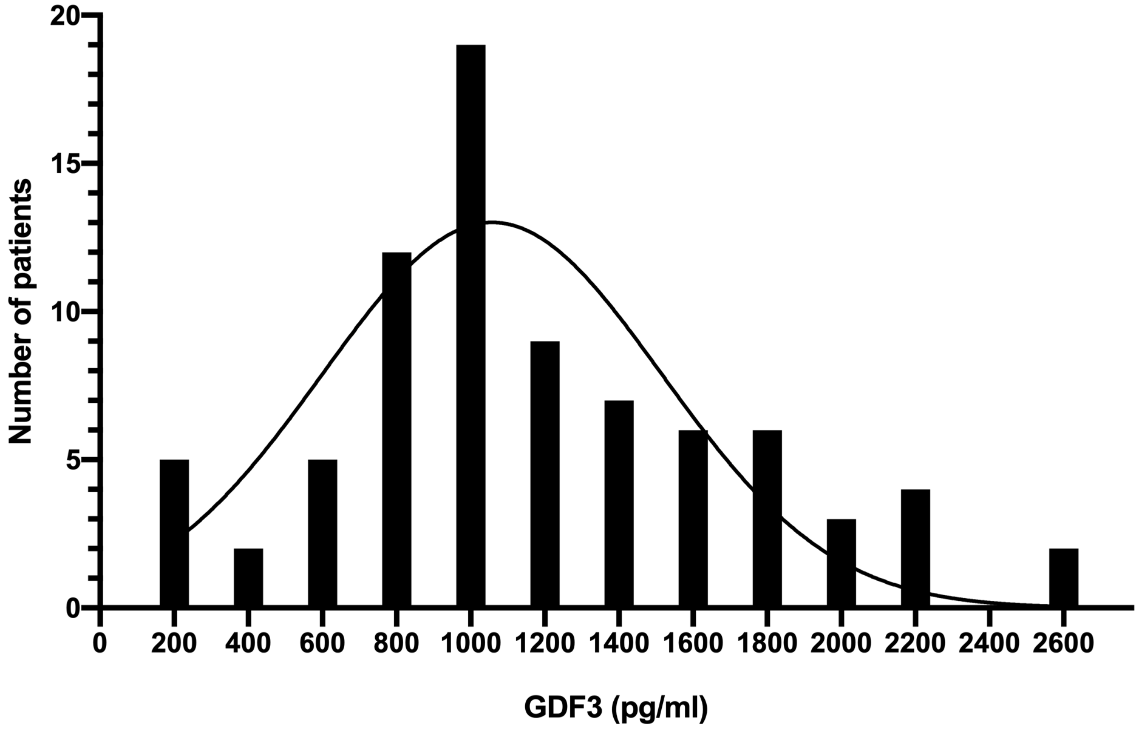

**Online Figure 4. Left ventricular end-diastolic volume indexed for body surface area measured on cardiac MRI 6 months after MI, according to GDF3 quartiles**. GDF3 was measured by ELISA in human plasmas collected at day 4±2 post-PCI and patients were classified into 4 quartiles according to GDF3 levels. Statistical significance was determined by one way ANOVA followed by comparison tests, *P < 0.05

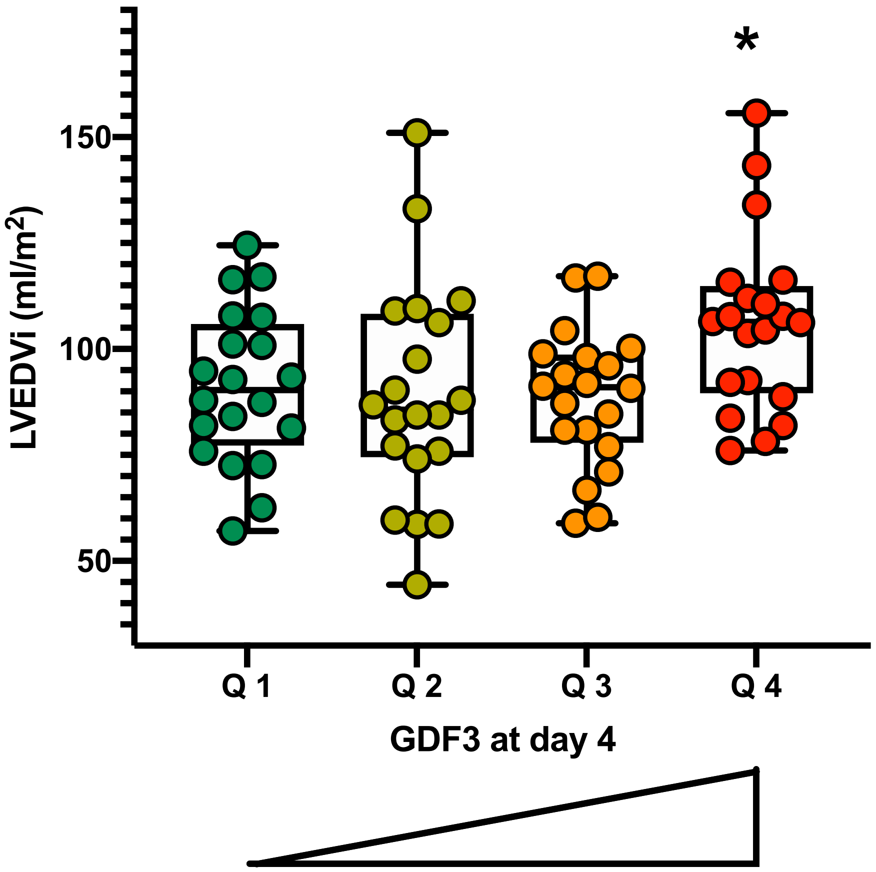


| | GDF3 levels (pg/mL) | | | |
|---|---|---|---|---|
| | **Quartile 1 [150-807]** | **Quartile 2 [825-1040]** | **Quartile 3 [1050-1510]** | **Quartile 4 [1550-2570]** |
| **LVEDVi** (ml/m$^2$) **Mean** | 91.0 | 89.1 | 88.3 | 105.9 |
| **SD** | 18.2 | 25.9 | 16.3 | 20.9 |

**Online Figure 5. Left ventricular ejection fraction measured on cardiac MRI 6 months after MI, according to GDF3 quartiles**. GDF3 was measured by ELISA in human plasmas collected at day 4±2 post-PCI and patients were classified into 4 quartiles according to GDF3 levels. Statistical significance was determined by one way ANOVA followed by comparison tests, *P < 0.05

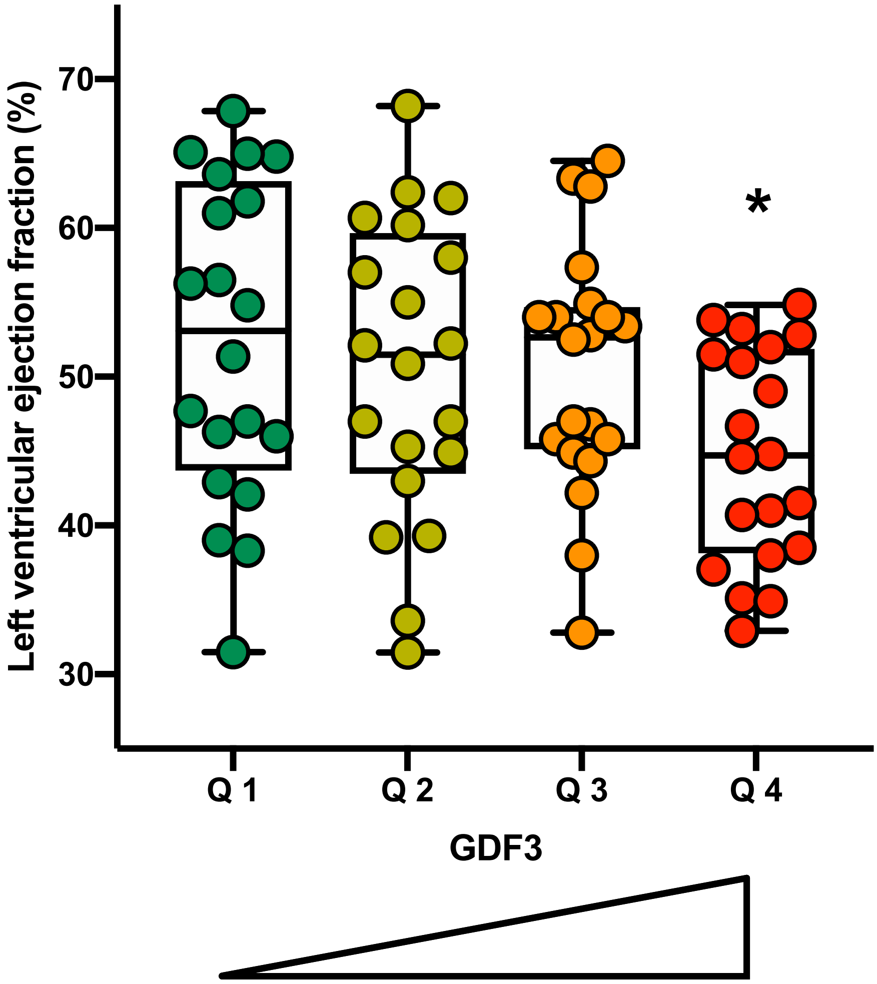


| | GDF3 levels (pg/mL) | | | |
|---|---|---|---|---|
| | **Quartile 1 [150-807]** | **Quartile 2 [825-1040]** | **Quartile 3 [1050-1510]** | **Quartile 4 [1550-2570]** |
| **LVEF** (%) **Mean** | 52.4 | 50.5 | 50.6 | 44.7 |
| **SD** | 10.7 | 10.1 | 8.3 | 7.2 |
